# Self-Collected Oral Fluid and Nasal Swabs Demonstrate Comparable Sensitivity to Clinician Collected Nasopharyngeal Swabs for Covid-19 Detection

**DOI:** 10.1101/2020.04.11.20062372

**Authors:** N Kojima, F Turner, V Slepnev, A Bacelar, L Deming, S Kodeboyina, JD Klausner

## Abstract

**Background:** Currently, there is a pandemic caused by the 2019 severe acute respiratory syndrome coronavirus 2 (SARS-CoV-2), which causes Covid-19. We wanted to compare specimen types and collection methods to explore if a simpler to collect specimen type could expand access to testing.

**Methods:** We recruited individuals recently tested for SARS-CoV-2 infection through a “drive-through” testing program. In participants’ homes, we assessed the performance of self-collected oral fluid swab specimens with and without clinician supervision, clinician-supervised self-collected mid-turbinate (nasal) swab specimens, and clinician-collected nasopharyngeal swab specimens. We tested specimens with a validated reverse transcription-quantitative polymerase chain reaction assay for the detection of SARS-CoV-2 and measured cycle threshold values. Symptom status and date of onset of symptoms was also recorded for each participant.

**Results:** We recruited 45 participants. The median age of study participant was 42 years old (Interquartile range, 31 to 52 years). Of the participants, 29 had at least one specimen test positive for SARS-CoV-2. Of those, 21 (73%) of 29 reported active symptoms. By specimen type and home-based collection method, clinician-supervised self-collected oral fluid swab specimens detected 26 (90%) of 29 infected individuals, clinician-supervised self-collected nasal swab specimens detected 23 (85%) of 27, clinician-collected posterior nasopharyngeal swab specimens detected 23 (79%) of 29, and unmonitored self-collected oral fluid swab specimens detected 19 (66%) of 29. Despite nasopharyngeal swabs being considered the gold standard, 4 participants tested negative by clinician-collected nasopharyngeal swab and positive by the 3 other specimen types. Additionally, false negative results by each sample type were seen to generally not overlap.

**Conclusions:** Supervised self-collected oral fluid and nasal swab specimens performed similarly to, if not better than clinician-collected nasopharyngeal swab specimens for the detection of SARS-CoV-2 infection. No sample type captured all SARS-CoV-2 infections, suggesting potential heterogeneity in the distribution of viral load in different parts of the respiratory tract between individuals. Supervised self-collection performed comparably to clinician collection and would allow for rapid expansion of testing capacity in the United States by reducing the need for trained healthcare workers, reducing exposure of healthcare workers, and reducing the amount of PPE (personal protective equipment) being used for testing during a critical shortage.

## MAIN TEXT

The 2019 severe acute respiratory syndrome coronavirus 2 (SARS-CoV-2), which causes Covid-19, was first detected in Wuhan, China in late 2019.^1^ On 20 January 2020, the first case of SARS-CoV-2 infection was reported in the United States.^2^ After more than 118,000 cases were detected in 114 countries with over 4,000 deaths, the World Health Organization declared that Covid-19 pandemic.^3^

The ideal specimen for the detection of SARS-CoV-2 is unknown. Currently, trained health care professionals and specialized collection devices are recommended for the collection of nasopharyngeal swab specimens.^4^ That requires staffing of health care workers, who could be performing other duties, and the use of personal protective equipment (PPE), during a severe shortage. Additionally, patients report discomfort during nasopharyngeal swab specimen collection, which may deter patients from being tested. The use of nasal swab and oral fluid specimens could potentially greatly increase health worker safety and the number of persons tested. We recruited participants recently tested for SARS-CoV-2 infection to assess differences in specimen types and collection methods for Covid-19 testing.

## Methods

We recruited participants that recently tested for Covid-19 at a Clinical Laboratory Improvement Amendments certified, high-complexity laboratory. The patient population and recruitment methods are described below.

### Testing population

We recruited non-hospitalized persons tested for Covid-19 in Los Angeles County, California, that included symptomatic adults older than age 65, those with a chronic disease, first responders, and law enforcement officers that may have been exposed to SARS-CoV-2. We aimed to recruit 30 persons that tested negative for Covid-19 and 30 persons that tested positive. Participants were contacted via telephone or email and provided with details of the study. Participants were given a study information sheet and gave verbal informed consent.

### Specimen collection methods

We aimed to study different supervised versus unsupervised, self-collected specimens. We obtained unsupervised self-collected oral fluid swab specimens, clinician-supervised self-collected oral fluid swab specimens, clinician-supervised self-collected mid-turbinate (nasal) swab specimens, and clinician-collected posterior nasopharyngeal swab specimens.

For the self-collected oral fluid swab specimens, we provided written instructions with the testing kit, which included a sterile swab and a tube with a ribonucleic acid storage media (DNA/RNA Shield™ solution, Zymo Research Corp., Irvine, CA, USA). Participants were instructed to cough deeply 3-5 times collecting any phlegm or secretions in their mouth, rub the swab on both cheeks, above and below the tongue, both gums, and on the hard palate for a total of 20 seconds to ensure the swab was saturated with oral fluid. Following that, participants were instructed to place the swab into the tube, secure the lid, invert the tube 3-5 times, and place the capped tube into a collection bag. For the clinician-supervised self-collected oral fluid swab specimens, the same instructions were provided and a clinician observed to provide real time feedback. We observed that without feedback, some unsupervised patients did not cough before self-collecting their sample.

For the clinician-supervised self-collected nasal swab specimen, a kit was provided that included a flocked swab (CLASSIQSwabs™, Copan Diagnostics, Murrieta, CA, USA) and the same collection fluid as described above. The participant was verbally instructed to insert the swab into one nostril to the depth of 3-4 cm, rotate the swab for 5 to 10 seconds, place the swab into the collection tube, invert the tube 3-5 times, and place the capped tube into a collection bag. Posterior nasopharyngeal swab specimens were collected by a clinician with the recommended medical technique using nasopharyngeal swabs (Becton, Dickinson and Company, Franklin Lakes, NJ, USA).^5^

### Surveying and sampling

We collected samples in private areas of participant homes. We collected symptoms data immediately prior to sampling. Sampling methods are detailed above. For each patient, all samples were collected within a 30-minute window. Samples were transported to the laboratory at ambient temperature for testing on the day of collection.

### Specimen extraction and testing

We processed samples from the specimen collection tubes. We lysed and extracted RNA from samples (RNA purification kit, Norgen Biotek Corp., Thorold, ON, Canada) using an automated instrument (Resolvex A200, Tecan Group Ltd., Zürich, Switzerland) on a 96-well plate (Norgen Biotek Corp., Thorold, ON, CA). We used a reverse transcription-quantitative polymerase chain reaction (RT-qPCR) assay that utilized a single color TaqMan probe with a modified version of the qualitative detection of Covid-19 (N1, N2 primer/probe assay) designed and validated by the Centers for Disease Control and Prevention (CDC), with the addition of N3 (Integrated DNA Technologies, Coralville, IA, USA).^6^ We recorded cycle threshold values for tests. We detected human Ribonuclease P RNA with an additional single color TaqMan assay, in a parallel reaction using an aliquot of the extracted participant specimen to serve as a control for specimen extraction, specimen adequacy, and RT-PCR inhibition. We ran samples on an RT-PCR System (CFX 96^™^ Touch RT-PCR Detection System or CFX 96^™^ Connect RT-PCR Detection System, Bio-Rad, Hercules, CA, USA).z

### Ethics statement

The Institutional Review Board of the University of California Los Angeles reviewed and approved the study (reference number 20-000545).

## Results

We recruited 45 participants. The median age of study participants was 42 years (Interquartile range [IQR], 31 to 52 years). Of the participants, 29 tested positive for SARS-CoV-2 viral RNA in at least one specimen. Of the participants, 23 (51%) of 45 participants reported active symptoms; 21 of those 23 tested positive.

Overall, we collected 180 samples from 45 participants. Of those specimens, one specimen was lost and two specimens had insufficient sample for laboratory analysis. Therefore, 177 specimens yielded results (Figure). Clinician-supervised oral fluid swab specimens detected 26 (90%) of 29 infected individuals, clinician-supervised nasal swab specimens detected 23 (85%) of 27, clinician-collected posterior nasopharyngeal swab specimens detected 23 (79%) of 29, and unsupervised self-collected oral fluid swab specimens detected 19 (66%) of 29. There was no difference in testing performance when comparing those with and without active symptoms.

**Figure.**
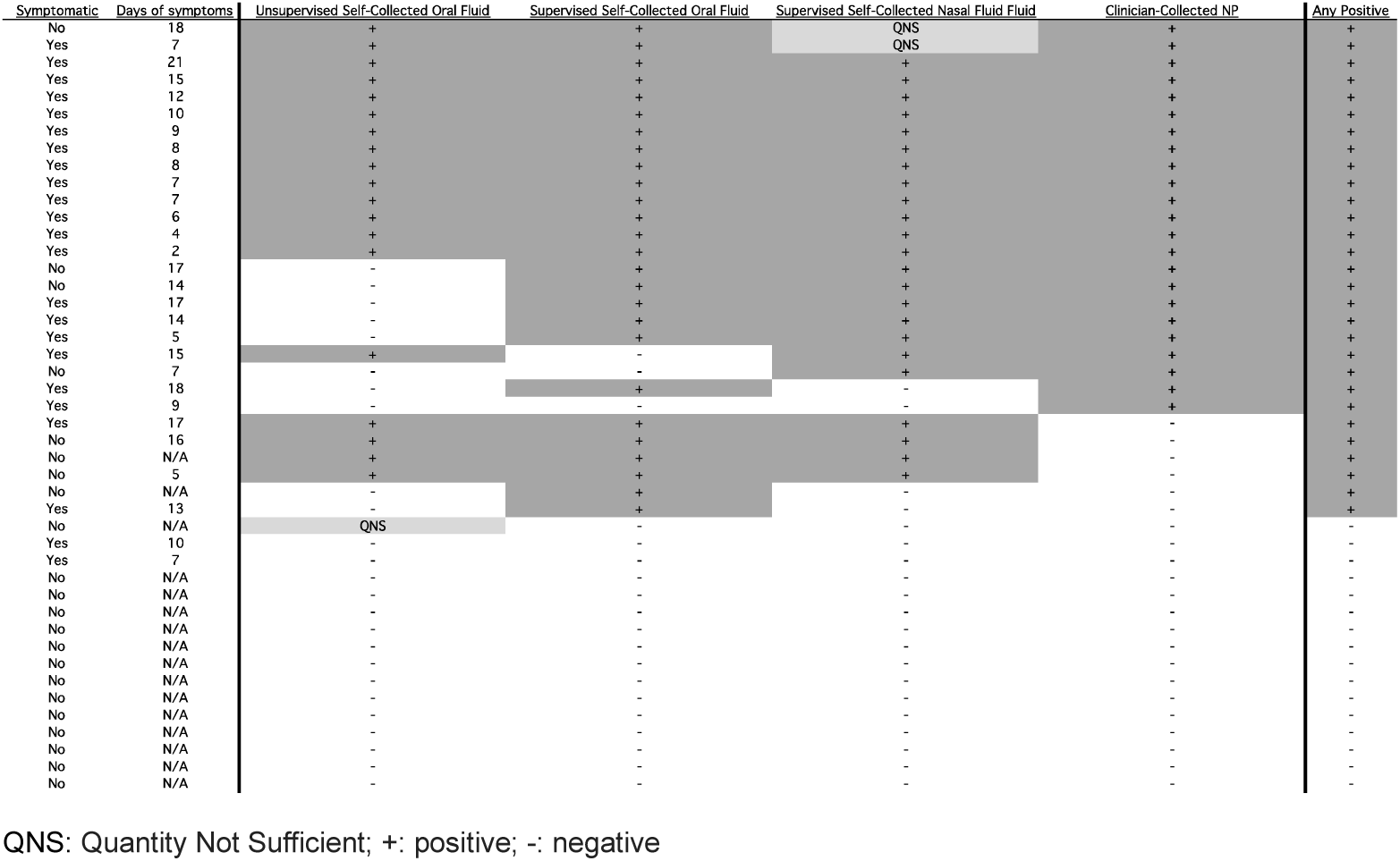
2019 severe acute respiratory syndrome coronavirus 2 infection detection in home-based self-collected clinician-supervised and unsupervised oral fluid swab specimens, clinician-supervised self-collected nasal turbinate swab specimens, clinician-collected posterior nasopharyngeal swab specimens, and pooled results with symptom status

When comparing cycle threshold values, clinician-collected posterior nasopharyngeal swab specimens had an average cycle threshold value of 15.96 (SD: 14.91), clinician-supervised self-collected nasal swab specimens had an average cycle threshold value of 19.25 (SD: 16.53), clinician-supervised self-collected oral fluid swab specimens had an average cycle threshold value of 19.72 (SD: 17.27), and unsupervised self-collected oral fluid swab specimens had an average cycle threshold value of 18.33 (SD: 18.02).

## Discussion

We found that self-collection of specimens for SARS-CoV-2 detection was feasible. No single specimen type detected all those with infection. Supervised home-collection of oral fluid and nasal secretions performed as well as, or better than, clinician-collected nasopharyngeal specimens. Unsupervised self-collection of oral fluid specimens might have performed worse in this study sample.

The CDC currently recommends the use of nasopharyngeal or oropharyngeal swab specimens either collected by a health care worker or self-collected mid-turbinate or anterior nares samples in symptomatic patients in a health care setting, including a supervised drive-through setting, if nasopharyngeal swab specimens are not available.^4^ A prior study reported that Covid-19 detection was similar among sputum and nasopharyngeal swabs specimens.^7^ That study noted that multiple anatomic site testing may improve the sensitivity and reduce false-negative test results.

There is an urgent need to validate reliable specimen collection methods for the detection of SARS-CoV-2 to increase access to safe and easy testing. We believe this is the first study demonstrating sample collection for SARS-CoV-2 infection testing in a home setting. Our findings support that clinician-supervised self-collected oral fluid and clinician-supervised self-collected nasal swab specimens for the detection of Covid-19 in the home setting are likely equivalent in sensitivity to clinician-collected posterior nasopharyngeal swab specimens. Further research on other supervision means such as video-based instructions or observation and feedback via telehealth is warranted.

In our sample, there were 6 cases of SARS-CoV-2 infection detected among oral fluid swab specimens, which were not detected in the clinician-collected nasopharyngeal swab specimens. There were also 3 cases of SARS-CoV-2 infection detected among nasopharyngeal specimens, not detected in oral fluid swab specimens. That suggests that testing any single anatomic site may miss some cases of Covid-19 infection, which is consistent with a prior study.^8^ While we did not find significant differences in cycle threshold values between groups, there was a trend toward swab specimens collected in the nasopharynx having lower cycle threshold values than swab specimens collected in the oropharynx, which correspond to higher viral loads in nasal or nasopharyngeal specimens.^9^

We found that unsupervised self-collected oral fluid swab specimens detected SARS-CoV-2 in fewer patients than other specimen types, and this discrepancy was unexpected. We observed that without feedback, some unsupervised participants did not cough before self-collecting their sample. Coughing was included as part of this specimen collection protocol. Laboratory studies and a case series have indicated that oral fluid collected after a participant coughs are reliable specimens.^10,11^ This study suggests that coughing may be a critical step when collecting oral fluid swab specimens for the detection of SARS-CoV-2.

Our report has several strengths. We were able to perform home-based specimen collection for Covid-19 testing. We studied multiple sample types and collection methods, including unsupervised self-collected specimens and clinician-supervised self-collected specimens. Clinician-collected nasopharyngeal specimens were collected in all patients for comparison. All samples were tested at a Clinical Laboratory Improvement Amendments certified, high-complexity laboratory with a validated Covid-19 assay.

However, our study had a limited sample size due to the current shortage of testing supplies. Our study was not designed to detect statistical differences between specimen types or collection methods. Given the urgency of obtaining results, recruitment took place over a short period.

## Conclusions

Supervised self-collected oral fluid and nasal swab specimens performed similarly to clinician-collected nasopharyngeal swab specimens for the detection of SARS-CoV-2. No sample type captured all infections. Supervised self-collected methods were feasible and could enable widespread access to testing by removing the need for a healthcare professional to collect each sample, reducing potential exposure for healthcare professionals and reducing the amount of PPE used for testing.

## Data Availability

Authors will share data with a reasonable data request.

## Acknowledgements

The authors want to acknowledge the staff of University of California Los Angeles, TKSL, Curative Inc. and Korva-Labs Inc.

